# Cortical microstructure and hemispheric specialization – a diffusion-imaging analysis in younger and older adults

**DOI:** 10.1101/2023.12.19.23300148

**Authors:** Paweł P. Wróbel, Hanna Braaß, Benedikt M. Frey, Marlene Bönstrup, Stephanie Guder, Lukas K. Frontzkowski, Jan F. Feldheim, Bastian Cheng, Yogesh Rathi, Ofer Pasternak, Götz Thomalla, Inga K. Koerte, Martha E. Shenton, Christian Gerloff, Fanny Quandt, Focko L. Higgen, Robert Schulz

**Author notes:** shared last authorship (Focko L. Higgen, Robert Schulz).

## Abstract

Characterizing cortical plasticity becomes increasingly important for identifying compensatory mechanisms and structural reserve in the aging population. While cortical thickness (CT) largely contributed to systems neuroscience, it incompletely informs about the underlying neuroplastic pathophysiology. In turn, microstructural characteristics may correspond to atrophy mechanisms in a more sensitive way, indicating a potentially necessary paradigm shift in neuroimaging. Fractional anisotropy (FA), a diffusion tensor imaging (DTI) measure, is inversely related to cortical histologic complexity. Axial (AD) and radial diffusivity (RD) are assumed to be linked to density of structures oriented perpendicular and parallel to cortical surface respectively. We hypothesized (1) that cortical DTI will reveal microstructural correlates for hemispheric specialization, particularly in the language and motor systems and (2) that lateralization of cortical DTI parameters will show an age effect, paralleling age-related changes in activation, especially in the prefrontal cortex. We re-analyzed data of healthy younger and older adult participants (n=91). DTI measures and CT were extracted from Destrieux atlas regions. Diffusion measures showed lateralization in specialized motor, language, visual, auditory, and inferior parietal cortices. Age-dependent increased lateralization was observed for DTI measures in the prefrontal, angular, superior temporal, and lateral occipital cortex. CT did not show any age-dependent alterations in lateralization. Our observations argue that cortical DTI is able to capture correlates of microstructural properties associated with functional specialization, resembling findings from histology. Age effects on diffusion measures in the integrative prefrontal and parietal areas may shed novel light on the atrophy-related plasticity in healthy aging.

**Significance statement:** Cortical thickness significantly contributed to systems neuroscience research related to cortical neuroplasticity. However, regarding the underlying cortical microstructure it remains an unspecific measure. With a strong lateralization in diffusion measures but not in thickness in specialized areas we demonstrate that cortical diffusion MRI is suitable to grasp microstructural features linked to specialization already described in histology literature. The findings in the lateralization of prefrontal and parietal cortical features may reflect age-related dynamic in cerebral activation. These results indicate the great potential of cortical diffusion tensor imaging in neuroscience and may even emphasize a necessary paradigm shift from the assessment of cortical macrostructure towards cortical microstructure for a better understanding of neuroplasticity and structure-function relationships in health and disease.

## Introduction

The concept of hemispheric lateralization is a fundamental cerebral characteristic in systems neuroscience. Alterations in lateralization have been shown to be altered depending on age with patterns of dedifferentiation (1-3). Findings that have been contextualized by empirical models like the hemispheric asymmetry reduction in older adults (HAROLD) model (4-9) can be observed primarily in the prefrontal cortex. However, cortical thickness (CT) appears to be limited for capturing hemispheric specialization and poorly elucidates the mechanisms of its modulations. CT inconsistently reflects hemispheric specialization of speech areas, which show a profound lateralization in functional studies (10-14). Moreover, one study on a large dataset did not show any significant relationships between CT lateralization and age (15).The assumption that age may lead to altered cortical lateralization appears reasonable as diffusion magnetic resonance imaging (MRI) data of the white matter have already evidenced HAROLD-like reduced microstructural lateralization, particularly in the prefrontal cortex (16, 17), coinciding with changes in glucose metabolism patterns (18). So far, the issue of cortical microscopic correlates of brain plasticity in healthy aging has not been systematically explored by diffusion MRI (19).

Over a century ago Brodmann (20), Ramón y Cajal (21) and Vogt and Vogt (22) advocated the presence of inter-regional diversity of cortical microstructure and thereby promoted fundamental concepts of specialization of the human brain (23). Over time, imaging studies of structure-function and structure-behaviour relationships have continuously formed an important area of research in fundamental and clinical neuroscience (24, 25). More recently, the aging population (26) has led to an even greater interest in a better understanding of age-dependent changes in cortical structure and its relation to functional and behavioural deterioration. The variability in cortical structural properties has been repeatedly related to behavioural aspects in healthy aging, and to deficits and trajectories of recovery in neurological disease. The analysis of CT, a surrogate of cortical macrostructure, dominates the field. However, the interpretation of CT and its integration with brain function or behaviour is complicated as it is a relatively rough anatomic measure. CT sums multiple components such as neurons, glia cells, extent of dendritic arborization, protein plaques and microangiopathy (27-31). In other words, CT can tell us about the size of a structure while its complexity remains elusive.

Diffusion tensor imaging (DTI) has revolutionized the microstructural analyses of the brain. Primarily, it has been applied to study white matter (32). However, technical progress, e.g., the introduction of free water correction (33), has increasingly empowered DTI to capture cortical microstructural properties. Comparative studies on DTI and histology found an overlap between tensors of both modalities, oriented orthogonally to the cortical surface due to columnar organization of neurons (34). Fractional anisotropy (FA), used to measure the extent of directed tensor orientation, i.e., a surrogate of complexity of cortical microstructure, was shown to decrease after birth as dendritic arborization in the cortex proceeds (35, 36). During lifetime, cortical FA was then reported to show an increase in frontal areas in young and middle-aged adults suggesting a decay in complexity of cortical microstructure (37), concurrent to synaptic pruning or neuronal degeneration. The understanding to what extent the novel cortical DTI may be capable to capture fundamental characteristics of cortical microanatomy is crucial to harness its potential.

Given the long-existing evidence for functional and microstructural hemispheric specialization and lateralization we first hypothesized that diffusion-based surrogate parameters of cortical microstructure will be capable to capture lateralized cortical features known from histology studies, particularly in the unimodal language and motor regions. Second, we hypothesized to detect age-dependent changes in the amount of hemispheric lateralization, which we expected, particularly, in heteromodal prefrontal cortices according to the HAROLD model. Following the interpretation of FA as a measure of cortical microstructural complexity (35, 36), we assumed that directed water diffusion along the cortical columns, probably representing the amount of neurons or afferent axons, would be best mapped by axial diffusivity (AD) whereas the diffusion along structures parallel to the cortex, such as dendrites and interneurons, would be best captured by radial diffusivity (RD) instead. To this end, we re-analyzed available imaging data of healthy younger and older adults from previously published studies (38-42) using diffusion imaging to obtain cortical FA, AD, and RD values for 74 anatomic regions per hemisphere and to infer the amount of hemispheric lateralization in these parameters. Lateralization patterns are reported for all participants, group comparisons and interaction analyses were performed to assess differences between age groups.

## Results

Based on the association between the diffusion signal and cortical structure we calculated FA, AD, RD, and CT for 74 Destrieux atlas regions from imaging data of 43 younger and 48 older healthy right-handed adults. The assessment of lateralization and its change with age was tested using linear mixed-effect models with side and age group covariates along their interaction, while adjusting for the nuisance variables sex, study and, in case of diffusion measures, also CT value. Figures 1 and 2 summarize the topography of regions exhibiting significant lateralization in cortical FA, AD, and RD, respectively. CT measures are shown for comparative purposes.

**Figure 1.**
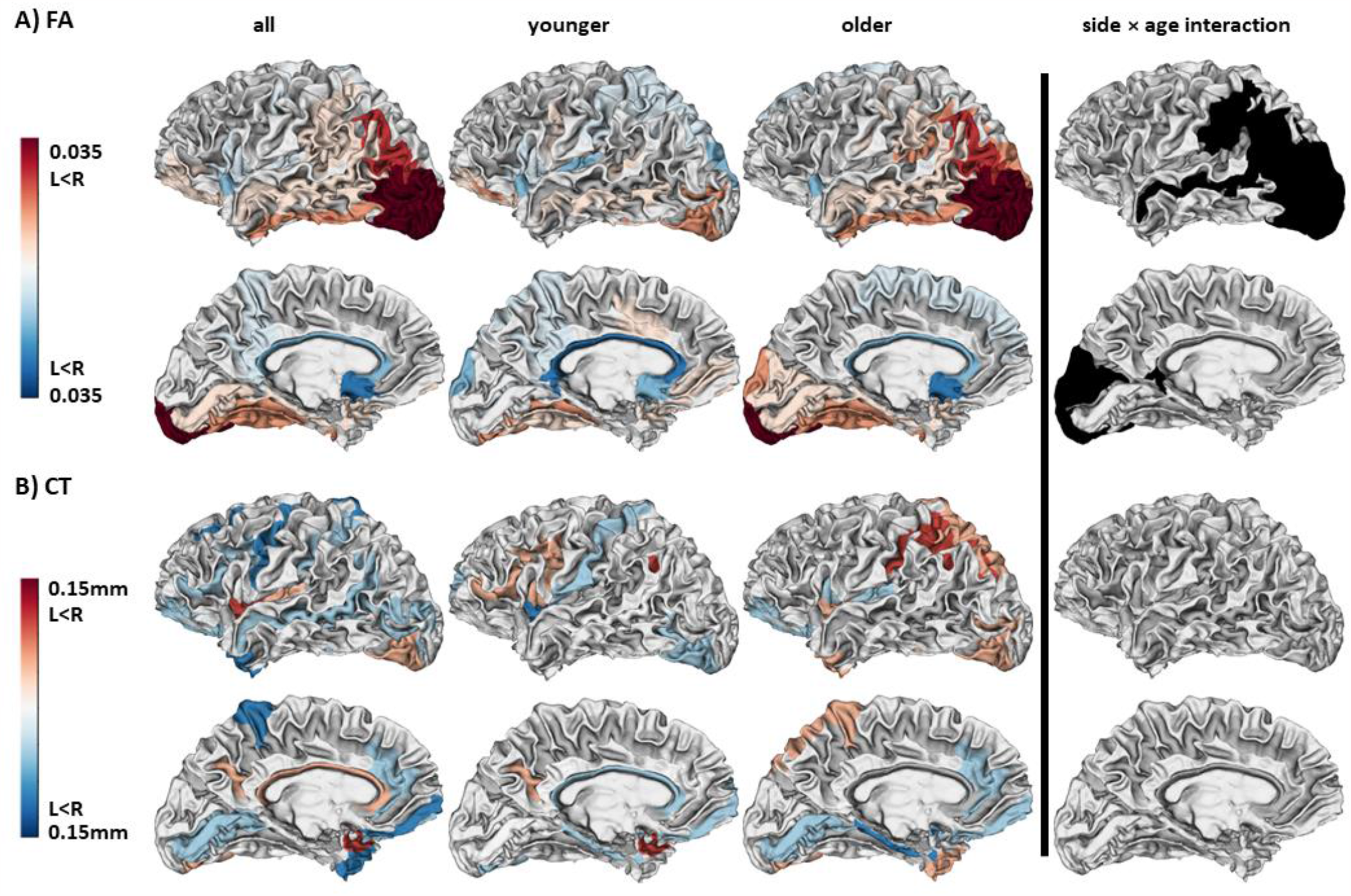
Lateralization of fractional anisotropy (FA) and cortical thickness (CT) (red - left < right values, blue – left > right values) in the combined group of participants (column 1), in younger (column 2) and older participants (column 3) as well as regions with significant interaction between the age group and lateralization. Displayed are the color-coded differences between hemispheres. Detailed results can be found in Supplementary Tables 1 and 2.

**Figure 2.**
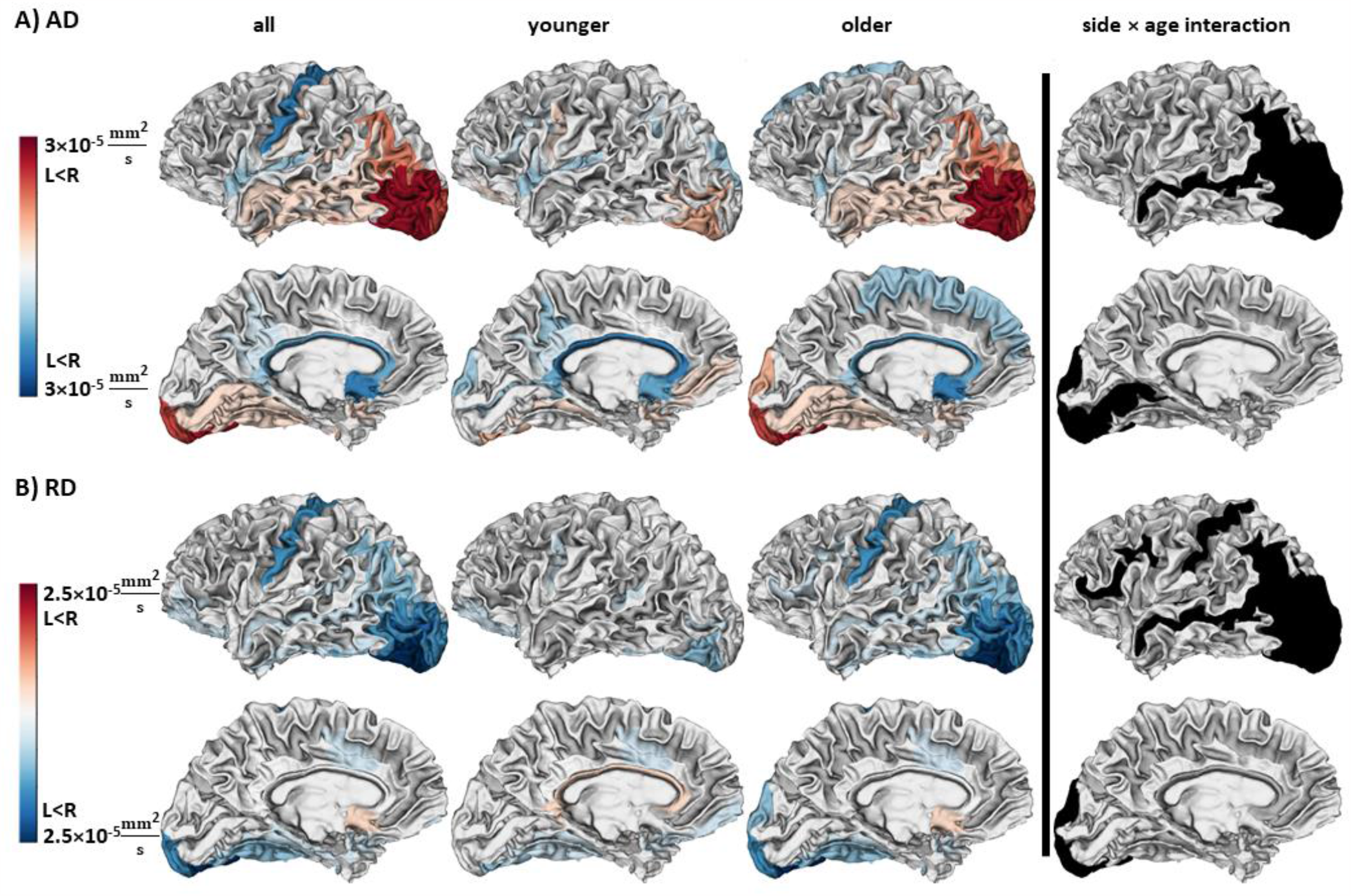
Lateralization of axial (AD) and radial diffusivity (RD) (red - left < right values, blue – left > right values) in the combined group of participants (column 1), in younger (column 2) and older participants (column 3) as well as regions with significant interaction between the age group and lateralization. Displayed are the color-coded differences between hemispheres. Detailed results can be found in Supplementary Tables 1 and 2.

In the combined group of younger and older adults, we found a lateralization with lower FA and AD in the dominant left hemisphere in the planum temporale and higher FA in the triangular portion of the dominant inferior frontal gyrus, i.e., areas contributing to Wernicke’s and Broca’s areas. In the planum temporale, findings were driven by lower left hemispheric AD in all groups, while RD was significantly greater in the left hemisphere only in younger adults. The lateralized FA in the triangular inferior frontal gyrus was determined by an isolated FA finding with greater left-hemispheric FA in younger adults. There were no significant interactions between lateralization and age group. CT did not show any significant lateralization in regions contributing to Wernicke’s and Broca’s areas.

A significant lateralization was also detected in the precentral gyrus, which comprises the primary motor cortex, one of the key areas of the human motor system. Here, we observed greater left hemispheric AD and RD but not FA in the combined group. Notably, within-group analyses for younger and older participants revealed that RD was significantly greater in the left hemisphere in older adults while AD showed a strong statistical trend towards a leftward lateralization (see Tables 1 and 2, Supporting Information). The younger adults did not show a significant lateralization in the precentral gyrus. Despite this numerical group difference, interaction models could not evidence any significant age effects for the precentral gyrus. There was no significant lateralization of CT in the precentral gyrus in any of the groups.

As examples for heteromodal brain areas, involved in motor and cognitive processes, hemispheric lateralization was also detected in the angular part of the inferior parietal lobe which showed a greater FA and AD in the right hemisphere. Greater RD was found in the left hemisphere in the combined group. The results were reproduced in the subgroup of older but not younger adults. The intraparietal sulcus displayed similar patterns of lateralization in DTI measures. In contrast to the language and motor areas, a statistically significant age effect was detected by interaction modeling in areas comprising the angular and intraparietal sulcal cortex. Given the clustering around the angular gyrus, detailed plots for diffusivity measures in both groups are provided in Supplementary Figure 3. Again, CT model yielded negative results also in these areas.

We additionally observed a lateralization of DTI measures in the calcarine sulcus, which comprises the primary visual cortex. However, greater left-hemispheric FA and AD were found only in younger adults.

We found age-dependent alterations in hemispheric microstructural asymmetry in prefrontal cortex in RD, but the greatest number of regions with significant interactions was clustered at the temporo-parieto-occipital (TPO) junction for all diffusion measures. There was no age-dependent lateralization in CT. As depicted in the images for sub-group analyses in Figures 1 and 2 (columns 1 – 3), RD revealed an increase in leftward asymmetry in older adults in the prefrontal cortex and the TPO cluster. AD and FA indicated greater values in the right hemisphere. A lateralization in these regions for younger was observed solely for FA in the intraparietal sulcus (Figure 1, column 2).

In the temporal lobe, analyses of superior sulcus in the combined group and the older adults indicated that FA and AD were associated with significantly lower left-hemispheric values. The RD showed relatively greater values in the left hemisphere. An interaction with age group was also detected for all three DTI measures. The middle temporal and fusiform gyri, the latter being associated with the visual word form area, showed lower leftward lateralization consistently across all groups for FA, while a consistent finding was observed for RD in the fusiform cortex.

## Discussion

The aim of this study was (1) to determine if diffusion-based surrogate parameters of cortical microstructure can detect histological traits associated with hemispheric specialization and (2) to evaluate if the amount of hemispheric specialization would show age-dependent changes. As the main findings we detected widespread lateralization of cortical microstructure, operationalized by the diffusion parameters FA, RD, and AD, respectively, particularly in the inferior frontal, precentral, superior, middle temporal, inferior parietal, and fusiform cortices. These cortices are not only part of language and motor networks, but also of occipital visual and posterior parietal multimodal integrational brain networks. Age-dependent increases in hemispheric lateralization were found primarily in the prefrontal cortex and in the temporo-parieto-occipital junction cortices.

### Hemispheric lateralization in structure and function

Based on comparative histology and DTI studies, we hypothesized to detect correlates of the structure-function relationship in the fundamental concept of hemispheric lateralization. Cortical FA was assessed as a surrogate parameter for cortical tissue complexity (34-36). Cortical AD was hypothesized to reflect the density of neurons, as these oblong structures facilitate diffusion along tensors’ main axis. Cortical RD was assumed to primarily correspond to structures facilitating diffusion oriented in parallel to the cortical surface, such as dendrites and interneurons. We hypothesized that hemispheric lateralization in these diffusion parameters would mirror hemispheric specialization, known from histology studies, particularly for the language or motor systems. We identified five regions exhibiting pronounced hemispheric specialization.

The first two regions were the planum temporale and the inferior frontal gyrus comprising Wernicke’s and Broca’s areas, core regions of the human language network. In the planum temporale we observed lower left-hemispheric FA, which could be induced by two different contributors. First, the accompanying reduction in left-hemispheric AD stands in line with existing histology findings: Greater intercolumnar gaps between neurons with less oblong neurons compared to the right-hemispheric homologue (43-45) can lead to smaller AD values and FA values on the left side. Second, denser and more widespread dendritic arbors in the left planum temporale (46-48), extending to a certain degree parallel to the cortical surface, might suggest a plausible link to the greater RD values observed in younger adults. Earlier studies proposed RD to be inversely linked with neuron density due to the influence of myelination (49). Broca’s area (assessed via the triangular inferior frontal cortex) has been characterized in a review by Keller *et al*. as poor in asymmetry on the macrostructural level and showing microstructural lateralization “more often than not” (10). While previous microscopy studies have reported multiple parameters to be lateralized in planum temporale, only a lower grey-level index (GLI) was observed for Broca’s area (50, 51), suggesting lower fraction of perikaryal structures, which could be based on greater amount of dendrites. One previous imaging study observed cortical microstructural lateralization for Wernicke’s area, but not for Broca’s area (19). In the present analysis, we found greater FA values in the left Broca’s area driven by data from younger adults. The link between cortical FA and the GLI remains speculative as the latter reflects the fraction of Nissl-stained ribosomal structures, and not the number of neurons per se as glia cells would also contribute to GLI. Such cell populations are less elongated, not as uniformly arranged as neurons in the cortex and highly different in cytomorphology. In contrast to the planum temporale, due to absence of positive AD or RD findings it remains elusive if the difference in FA is driven by differences in complexity or in density of axonal structures.

As a third region, greater AD and RD were measured in the left, dominant hemispheric precentral gyrus which comprises the primary motor cortex. It was striking not to observe a lateralization of FA in the precentral area. However, given the nature of FA as a ratio, even a more complex tissue may still show unchanged FA, if the amount of diffusivity along neuronal columns remains relatively equal compared to the diffusion in the plane perpendicular to neuronal column’s axis. This observation compels to evaluating cortical diffusion imaging in the light of diffusivity measures of AD and RD to allow more precise statements on the microstructure. For instance, greater AD in the left and dominant motor cortex indicates greater diffusivity perpendicular to cortical surface and could therefore corresponds to the neuronal density, which was shown to be greater in the left primary motor cortex (52). In turn, a greater left RD (diffusivity parallel to the cortical surface), significant only in older adults, could point towards a greater amount of dendritic or inter-neuronal structures.

Fourth, lateralization could be found beyond the eloquent areas of language and motor networks. Lower FA, AD, and greater RD in the left-hemispheric inferior parietal cortex and the temporo-parieto-occipital junction could correspond to specialization of the parietal cortex in higher-order integrative functions that was observed in imaging and stimulation studies (8, 53), while ischemic lesions affecting right inferior parietal cortices have been linked with neglect and apraxia (54, 55). Strikingly, one would expect greater AD and RD, in particular when considering the specialization, in the right hemisphere as these could represent greater neuronal count and dendritic arborization or interneuronal density in the specialized area. However, as the visualized data for the angular gyrus from both subgroups suggest (see Supplementary Figure 3), the lateralization of AD and RD becomes apparent in late adulthood. Concurrently, the absolute level of AD is greater and the absolute level of RD is lower in the older group, which could correspond to a greater axonal density and lower dendritic axonal complexity, induced by reduction of non–axonal structures along a constant amount of neurons. This is in line with earlier descriptions in review literature (56).

Fifth, the observed lateralization of the visual system in younger adults was a striking finding. Even though there are single reports on interhemispheric differences in activity of the calcarine cortex (9), the literature on this topic is sparse compared to data on asymmetry in motor and language function. Future studies will have to confirm this result.

Taken together, the number, the topography, and the side specificity in lateralization of cortical DTI measures encourage viewing these as a promising surrogate for functional hemispheric specialization. For instance, the RD findings appear to resemble histological findings on dendritic arborization in the planum temporale (43, 47) while AD lateralization corresponds with findings on neuronal/axonal density in the planum temporale (43, 57), the fusiform cortex (58) and the inferior parietal lobe (59). This agreement of lateralized cortical DTI metrics and classical histology supports the view that DTI is able to capture cortical microstructural traits which remain hidden for CT based morphometric analyses in the motor or language network. The absence of a positive finding for CT compared to the results of Kong *et al*. could rely on the larger sample size (n > 17000) which indicates that surrogates for microstructure could be not only more specific but also more sensitive (15).

### Influence of age on microstructural hemispheric specialization

Our second hypothesis was that cortical DTI metrics are able to detect age-dependent changes in hemispheric lateralization. Multiple functional MRI and EEG studies have shown alterations in hemispheric activation with age, particularly in prefrontal cortices which has been contextualized with theoretical models such as the HAROLD model (4, 60). Reduction in hemispheric asymmetry of functional activity in older adults (i.e. the HAROLD model) has been argued to reflect compensatory mechanisms to overcome a decline in cortical integrative function (9). In this study, the inferior frontal sulcal and the inferior parietal cortex, two heteromodal integrative areas, displayed an age-dependent increase in lateralization due to a stronger right-hemispheric lateralization in absolute measures (Figures 1 and 2 – fourth column, Supplementary Figure 3). Such dynamic of lateralization towards the opposite hemisphere is highly suggestive for a reorganization that might be linked to compensatory activity. From a clinical perspective, the distribution of significant findings in the inferior parietal cortex would be in line with the assumption that cortical aging and degenerative disorders primarily affect higher cognitive and integrational processes due to subtle reorganization rather than the function of primary areas (61). Even though the second hypothesis appears to be confirmed considering the finding of lateralized RD in prefrontal microstructure, the results’ distribution (Figures 1 and 2 - fourth column), is highly suggestive that age-dependent organization is more strongly pronounced in posterior brain regions. Besides the HAROLD model, aging-related processes have been proposed to show a posterior to anterior shift, contextualized by the PASA model (62). The findings in the TPO cluster in this study seem to support the assumption of this shift, which has already been discussed by Rathi *et al*. in analyses on DTI measures on a gross, lobar level (37).

Of note, the focus on structure in this study allows only speculative judgements on the underlying mechanisms. The morphology of the aging cortex has been characterized by distinct and focal changes in dendritic branching rather than by extensive neuronal loss (56), which would be primarily reflected by a reduction of RD and to lesser extent also in FA. Such reorganization would unlikely cause reorganization measured by AD. However, an altered diffusion along the neuronal columns may be also explained by altered afferent fibers resulting from white matter lesions leading to a disconnection syndromes due to Wallerian degeneration (16, 17). An earlier study of our group showed that better performing older adults show a greater white matter integrity, operationalized by regional FA, compared to low performing peers (39). Again, cortical DTI measures, grasping such nuances based on single-shell imaging, appear to be superior to CT based measurements (15).

### Implications

Cortical DTI with apparent advantages over CT bears a great potential for future clinical research questions. Current implications primarily focus on diagnostics. The observed relationship between AD and neuronal density as well as between RD and dendritic arborization may encourage researchers to further promote the usage of cortical DTI to address potential cortical microstructural alterations in neurological diseases (63). Furthermore, a better characterization of temporal and spatial aspects of neuroplasticity could reveal targets for stimulation techniques, secondarily improving their efficacy and efficiency.

### Methodical limitations

There are several important limitations to note: First, five different studies were re-analyzed in this work. We controlled our models for study category, despite the acquisition on the same scanner. The inconsistent T1-data resolution in two studies is unlikely to confound the results as it had to be downgraded to the resolution of the diffusion data prior to DTI measure estimation. Second, the study does not allow a lifespan analysis due to the cross-sectional character, indicating the necessity to verify our observations in a longitudinal dataset. Third, beside absent information on education and socioeconomic background the behavioral data of some studies lacked standardized handedness testing such as the Edinburgh Handedness Inventory. Instead, the participants had to declare their handedness, which could potentially lead to inclusion of participants trained to right-handedness. Fourth, the diffusion image resolution was 2mm, this might be rather lower when aiming to assess cortical microstructure. However, given the strong differences observed in a conservative analysis and standing in line with existing histology reports, we consider the results as robust. Nevertheless, cortical DTI analyses should be further improved using high-resolution protocols in the future. Fifth, due to existing but still sparse literature it is challenging to compare the absolute DTI measures results to other studies. Even though we could reproduce insular cortex values known from previous studies (64), future work must validate our results. Differences in scanners, resolution of MRI datasets and different approaches of preprocessing might influence cortical DTI metrics. However, we assume that measuring lateralization, in which every participant serves as his or her own control, sufficiently accounts for this limitation.

### Conclusion

Cortical DTI measures unambiguously grasped microstructural hemispheric lateralization in core areas known for asymmetry in function such as the motor and language networks, with FA showing different degree of variation between areas of higher and lower integrative function. This work further demonstrated age-related reorganization processes in integrative heteromodal, secondary cortices, which corresponds to findings from studies on cerebral activity. From a methodical perspective the present findings indicate, that (1) cortical DTI grasps not only myelinated structures within white matter but also the complexity from non-myelinated tissue in the cortical gray matter and (2) the diffusivity measures AD and RD may reflect specific histoarchitectural traits, such as neuronal density and dendritic arborization, being therefore compulsory for interpretation of FA in the cerebral cortex.

## Materials and methods

### Participants Data

Imaging and clinical data were screened from five individual datasets contributing to studies which have previously been published by our laboratory (38-42). Datasets of healthy right-handed participants within two age-ranges (18-30 and 60-87 years) with sufficient data quality were included in this secondary analysis. In total, 91 of initially 123 datasets were analyzed (number of included datasets from each study - Study A (38): 14 of 23 [older adults], Study B (39): 39 of 50 [19 younger and 20 older adults], Study C (40): 18 of 20 [younger adults], Study D (41): 8 of 13 [6 younger and 2 older adults], Study E (42): 12 of 17 [older adults]). The exclusions were based on age beyond the range of interest and data quality. In sum, age groups consisted of 43 younger (mean 24.4 years, standard deviation [SD] 2.5, range 18-30, 26 female) and 48 older participants (mean 71.7 years, SD 6.0, range 60-87, 22 female). Brain imaging for all participants had been conducted on the same MRI scanner. The original studies were conducted following the Declaration of Helsinki and approved by the local ethics committee of the Medical Association of Hamburg. All participants gave written informed consent.

### Brain Imaging and Processing

All participants were scanned on a 3 Tesla Siemens Skyra Scanner (Siemens Healthineers, Erlangen, Germany) with a 32-channel head coil. T1-weighted imaging was based on a magnetization-prepared, rapid acquisition gradient-echo sequence (MPRAGE) (repetition time (TR) = 2.500ms, echo time (TE) = 2.12ms, 256 slices with a field of view (FOV) = 240×192mm, interslice distance of 0.94mm and an in-plane resolution range between 0.94x0.94mm and 0.83×0.83mm). DTI datasets were consistent in imaging parameters, with 75 slices in 64 non-collinear gradient directions with a b-value of 1500s/mm^2^ as well as one b_0_ image (FOV = 208 × 256 mm^2^, flip angle = 90°, TE = 82ms, TR = 10000ms and voxel size of 2×2×2mm). Each individual dataset underwent manual quality check excluding datasets with motion artifacts, for instance ringing, ghosting or tissue cuts, leading to insufficient reconstruction of grey and white matter boundary, and datasets with cerebral tissue outside the FOV.

T1-weighted images were segmented into 74 cortical anatomic regions per hemisphere from the Destrieux atlas (65) using the FreeSurfer based *recon-all* tool (version 6.0.1) (66) yielding CT values and individual labels for identification of regions in DTI data. The data was post-processed and registered to the individual pre-processed DTI using Advanced Normalization Tools (ANTs 2.3.4) (67) while adjusting the resolution to 2x2x2mm. DTI datasets were corrected for eddy currents using MRtrix3 (3.0.2) tools based on the FSL (6.0.1) correction method (68, 69). Distortion correction was applied by registering the b_0_-weighted diffusion image to the T1-weighted image using ANTs, MRTrix3 and FSL. For free water (FW) correction of the cortical DTI signals, a custom written MATLAB script was used (ran on Matlab R2020a, The Mathworks, Natick, MA, USA). In brief, a bi-tensor model with a fixed diffusivity for FW and a second representing water diffusion in presence of tissue membranes (33) was fitted and mean FA, AD and RD values were derived from FW-corrected tensor eigenvalues for each cortical area.

### Statistical Analysis

Statistics were performed using R software version 4.0.2 (70). Linear mixed-effects models with repeated measures were fitted to describe the extent of label-wise hemispheric lateralization in cortical microstructure. Specifically, in separate models for FA, AD, RD, and CT in all participants (N=91), mean absolute values were treated as the dependent variable (DV), SIDE (left, right) was treated as the independent variable of interest. Target effects were adjusted in a multivariate approach for the nuisance variables AGE_GROUP, AGE_GROUP × SIDE interaction, STUDY, SEX, as well as CT in models for diffusion measures. The necessity to adjust for CT is explained in the supplementary text 1, see Supporting Information. The amount of hemispheric lateralization is based on the coefficient SIDE (mean difference between hemispheres), its statistical significance (*P*_SIDE_) is fully corrected for multiple comparisons using the false discovery rate (FDR) method for 74 Destrieux atlas regions per DTI parameter. The age-dependent change of lateralization is based on the coefficient AGE_GROUP × SIDE, its statistical significance (*P*_AGE_GROUP × SIDE_) was corrected in the same manner as the SIDE effect. To specifically address age effects, separate models were also fitted for the younger (N=43) and older (N=48) participants separately, with AGE as covariate. Statistical significance was assumed at corrected *P*_FDR_<0.05. Brain illustrations were generated using the *fsbrain* package in R.

## Data Availability

Data availability is limited by the approval of Ethics Committee

## Acknowledgments and funding

This work was supported by the German Research Foundation (DFG) and the National Science Foundation of China (NSFC) in project Crossmodal Learning, TRR-169/A3 (C.G.), the Deutsche Forschungsgemeinschaft (DFG, German Research Foundation) – SFB936-178316478 - C1 (C.G) & C2 (G.T., B.C.), Werner Otto Stiftung (4/90, R.S.), Else Kröner-Fresenius-Stiftung (grant numbers 2020_EKES.16, R.S.).

## Notes

### Competing Interest Statement

The authors have declared no competing interest.

### Author Declarations

Ethics committee of Physician Chamber of Hamburg gave ethical approval for this work.

