## Supplementary material for "Cortical microstructure and hemispheric specialization – a diffusion-imaging analysis in younger and older adults": SI

**This PDF file includes:**

Supporting text
Figures S1 to S3
Tables S1 to S2

### Supporting Text

#### Relationship between DTI measures and CT

Despite FW correction, cortical DTI measures can be also influenced by partial volume effects (PVE) from white matter diffusion signal. PVE may be dependent on CT, which may differ from the diffusion imaging voxel size, thus increasing FA in areas with low CT. The interpretation of results without CT adjusting remains unclear. Given the lack of literature on this issue a corresponding investigation was necessary prior to hypothesis testing. We assumed that cortical DTI measures are primarily linked to microstructure and its interregional variability, but not explained or driven by CT variability. For each DTI measure we computed a linear mixed-effects models with repeated measures based on left hemisphere data (minimizing specialization effects) of the younger participants group (minimizing aging/atrophy effects). The fixed effects included CT (continuous), REGION (categorical, 74 levels) and nuisance variables AGE (continuous), SEX and STUDY (categorical). participant ID was treated as a random effect. DTI parameters were considered as the dependent variable. CT and REGION were significantly associated with all three DTI measures (CT: $P = 0.024$, $P = 0.003$ and $P = 0.884$ for FA, AD, RD, respectively, REGION: all $P < 2\times10^{-16}$). Explained variance in the DTI measures was up to 0.67 in pseudo-$R^2$ for FA (0.62 for AD, 0.58 for RD). We hypothesized that the effect of REGION would dominate the variance explained. Consecutively, we omitted REGION from the modelling. The association between CT and the DTI measures remained significant, however, the association was rather weak given a critical drop in explained variance (pseudo-$R^2$, FA 0.06, AD 0.05, RD 0.08, see Supplementary Figures 1 and 2). Nevertheless, in order to exclude any confounding effect of CT variability onto DTI measures we opted to include CT as a nuisance variable in the subsequent statistical analyses.

**Fig. S1.**

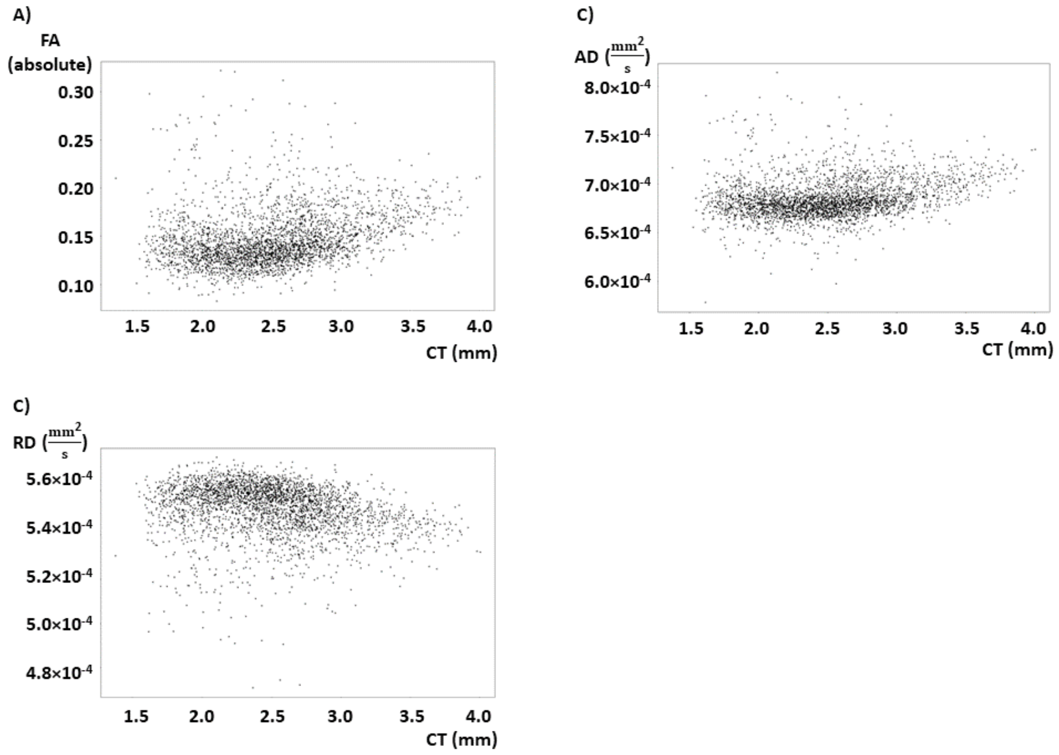


**Plotted raw diffusion measures and cortical thickness (CT) for 74 Destrieux regions of each 43 datasets from the subgroup of younger adult participants.**
A) Fractional anisotropy (FA) and CT. B) Axial diffusivity (AD) and CT. C) Radial diffusivity (RD) and CT.

**Fig. S2.**

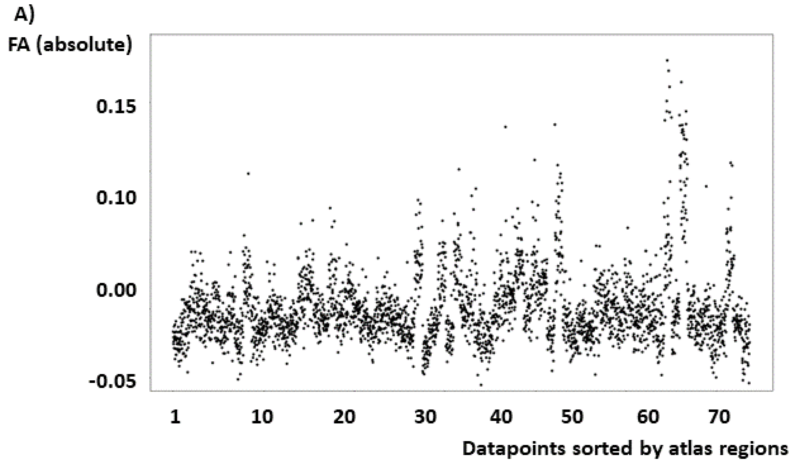


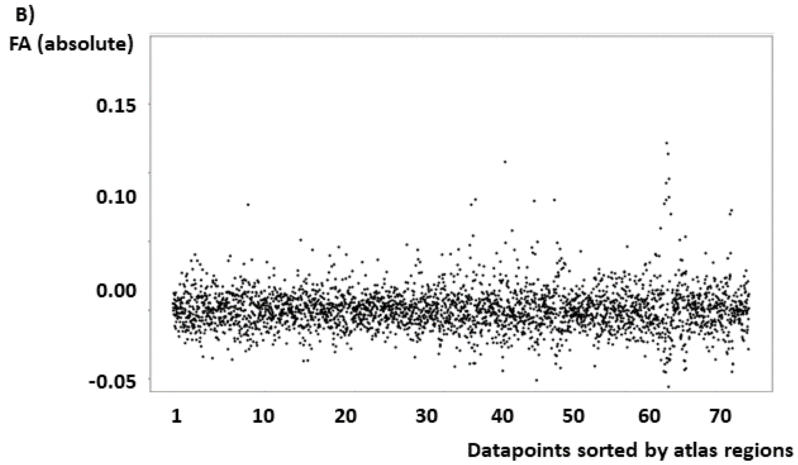


**Plotted residuals from modelling of fractional anisotropy (FA) from CT in a repeated measures model for a model without and with an interaction with the variable REGION.**
A) Residuals from the linear mixed-effects model [FA ~ CT + AGE + SEX + STUDY + (1|ID)] presented for dataset repeated for each region from the Destrieux atlas.
B) Residuals from the linear mixed-effects model [FA ~ CT × REGION + AGE + SEX + STUDY + (1|ID)] presented for dataset repeated for each region from the Destrieux atlas.

**Fig. S3.**

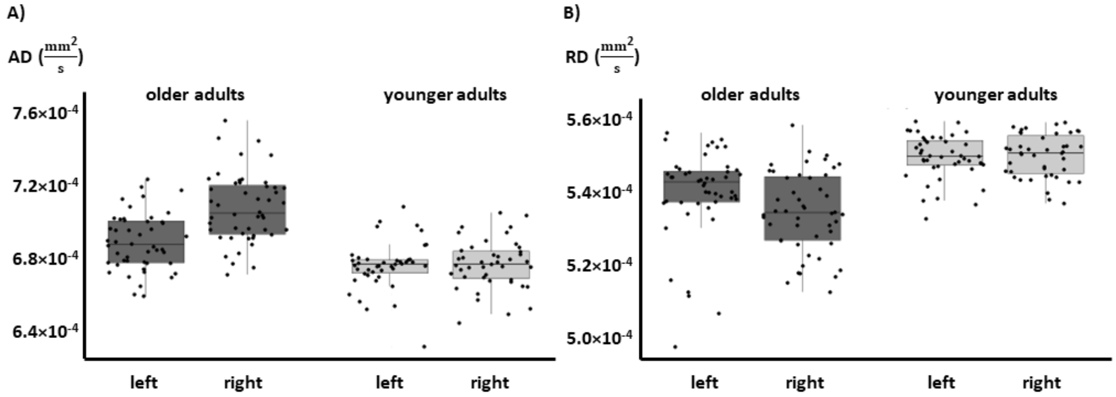


**Lateralization of A) axial (AD) and B) radial diffusivity (RD) in both age groups for the angular gyrus in the inferior parietal lobe.**

A post-hoc pairwise Tukey's from the original model indicate significant differences in diffusivity between age groups. All measures are given in mm²/s. AD: right hemisphere: $P < 0.001$ (estimated mean older adults: $7.12×10^{-4}$ [CI: $7.07×10^{-4}$ – $7.18×10^{-4}$], younger adults $6.75×10^{-4}$ [CI: $6.70×10^{-4}$ – $6.81×10^{-4}$]), left hemisphere $P = 0.001$ (estimated mean older adults: $6.94×10^{-4}$ [CI: $6.88×10^{-4}$ – $6.99×10^{-4}$], younger adults $6.75×10^{-4}$ [CI: $6.70×10^{-4}$ – $6.81×10^{-4}$]),. RD: right hemisphere: $P < 0.001$ (estimated mean older adults: $5.33×10^{-4}$ [CI: $5.29×10^{-4}$ – $5.37×10^{-4}$], younger adults $5.51×10^{-4}$ [CI: $5.48×10^{-4}$ – $5.55×10^{-4}$]), left hemisphere $P = 0.001$ (estimated mean older adults: $5.39×10^{-4}$ [CI: $5.35×10^{-4}$ – $5.42×10^{-4}$], younger adults $5.51×10^{-4}$ [CI: $5.48×10^{-4}$ – $5.55×10^{-4}$]).

### Table S1. Results overview

| | AD | | FA | | RD | | CT | | Significance level of AGE GROUP × SIDE interaction | | | |
|---|---|---|---|---|---|---|---|---|---|---|---|---|
| Region | coefficient | $P_{FDR}$ | coefficient | $P_{FDR}$ | coefficient | $P_{FDR}$ | coefficient | $P_{FDR}$ | $P_{FDR}$ AD | $P_{FDR}$ FA | $P_{FDR}$ RD | $P_{FDR}$ CT |
| G&S_cingul-Ant | -1.17E-06 | 6.31E-01 | -2.46E-03 | 6.21E-01 | 5.70E-07 | 7.89E-01 | -5.83E-03 | 2.37E-05 | 5.67E-01 | 3.44E-01 | 5.29E-01 | 8.76E-01 |
| G&S_cingul-Mid-Ant | 1.32E-06 | 2.12E-02 | 1.39E-03 | 5.77E-02 | -1.63E-06 | 2.03E-03 | -1.42E-02 | 9.41E-01 | 3.99E-01 | 3.44E-01 | 7.83E-01 | 5.71E-01 |
| G&S_cingul-Mid-Post | -1.41E-06 | 3.44E-01 | 1.12E-03 | 9.25E-01 | -8.59E-07 | 7.14E-01 | -4.36E-02 | 3.01E-01 | 9.40E-01 | 5.70E-01 | 5.62E-01 | 5.81E-01 |
| G&S_frontomargin | -2.29E-06 | 5.96E-01 | -1.19E-03 | 7.32E-01 | -1.18E-06 | 5.14E-01 | -1.59E-02 | 8.99E-01 | 9.67E-01 | 9.90E-01 | 8.02E-01 | 7.26E-01 |
| G&S_occipital_inf | 3.52E-05 | 1.38E-11 | 5.27E-02 | 4.69E-18 | -2.42E-05 | 1.41E-11 | 9.45E-02 | 4.57E-09 | 1.28E-03 | 5.07E-06 | 6.51E-05 | 6.54E-01 |
| G&S_paracentral | -8.17E-06 | 1.36E-01 | -3.59E-03 | 6.86E-03 | -5.56E-06 | 9.58E-01 | -6.10E-02 | 4.61E-02 | 4.09E-01 | 4.17E-01 | 1.88E-01 | 3.38E-01 |
| G&S_subcentral | 2.75E-06 | 1.55E-01 | 1.89E-03 | 6.75E-01 | 1.60E-06 | 1.47E-01 | -6.60E-02 | 4.54E-01 | 3.14E-01 | 4.17E-01 | 5.25E-01 | 1.69E-01 |
| G&S_transv_frontopol | 5.50E-06 | 5.17E-01 | -6.91E-03 | 2.53E-01 | 1.11E-05 | 1.36E-01 | 2.13E-02 | 8.68E-02 | 9.80E-01 | 3.06E-01 | 5.62E-01 | 4.50E-01 |
| G_cingul-Post-dorsal | -1.84E-06 | 9.99E-03 | -2.05E-03 | 1.31E-02 | 7.68E-07 | 1.54E-02 | 1.52E-02 | 5.58E-02 | 6.00E-01 | 3.01E-01 | 4.34E-01 | 5.71E-01 |
| G_cingul-Post-ventral | -6.18E-06 | 3.49E-08 | -7.13E-03 | 3.79E-07 | 1.93E-06 | 2.28E-04 | -6.61E-02 | 1.18E-01 | 1.01E-01 | 3.47E-02 | 1.81E-01 | 5.71E-01 |
| G_cuneus | 3.38E-06 | 8.88E-01 | 6.87E-03 | 6.59E-01 | -2.70E-07 | 3.73E-01 | -3.19E-02 | 1.18E-01 | 8.26E-02 | 1.83E-02 | 2.79E-01 | 8.29E-01 |
| G_front_inf-Opercular | 2.37E-06 | 6.24E-01 | -1.46E-03 | 6.59E-01 | 3.02E-06 | 5.34E-01 | -1.87E-02 | 3.13E-01 | 7.72E-01 | 7.48E-01 | 4.88E-01 | 3.38E-01 |
| G_front_inf-Orbital | -2.74E-06 | 7.69E-01 | -1.15E-03 | 9.25E-01 | 3.15E-08 | 9.89E-01 | 2.17E-02 | 6.53E-01 | 7.51E-01 | 8.10E-01 | 9.87E-01 | 7.26E-01 |
| G_front_inf-Triangul | -2.11E-06 | 3.87E-01 | -2.14E-03 | 1.91E-02 | -1.45E-06 | 9.14E-01 | -1.31E-02 | 5.15E-02 | 9.67E-01 | 3.01E-01 | 4.34E-01 | 9.85E-01 |
| G_front_middle | -1.74E-06 | 7.92E-01 | -7.81E-04 | 6.75E-01 | -1.40E-06 | 8.56E-01 | -3.98E-02 | 1.33E-01 | 7.50E-01 | 9.59E-01 | 6.59E-01 | 3.85E-01 |
| G_front_sup | -7.82E-06 | 1.36E-01 | -3.38E-03 | 5.76E-02 | -4.87E-06 | 4.18E-01 | -3.43E-02 | 2.60E-01 | 1.80E-01 | 2.34E-01 | 3.09E-01 | 6.54E-01 |
| G_Ins_lg_and_S_cent_ins | -5.86E-06 | 3.18E-04 | -6.49E-03 | 1.39E-03 | 3.11E-06 | 2.28E-04 | 8.31E-02 | 1.01E-05 | 7.34E-01 | 5.16E-01 | 7.08E-01 | 3.38E-01 |
| G_insular_short | -5.11E-06 | 1.27E-03 | -7.54E-03 | 3.56E-03 | 2.20E-06 | 5.49E-03 | 3.08E-01 | 6.89E-05 | 9.40E-01 | 8.47E-01 | 8.17E-01 | 7.28E-01 |
| G_oc-temp_lat-fusifor | 9.06E-06 | 9.99E-07 | 1.38E-02 | 1.55E-07 | -6.50E-06 | 1.35E-08 | 2.94E-02 | 2.04E-02 | 8.05E-02 | 8.26E-02 | 1.88E-01 | 5.10E-01 |
| G_oc-temp_med-Lingual | 9.52E-06 | 9.99E-07 | 5.30E-03 | 1.63E-02 | 2.94E-06 | 6.24E-02 | -2.16E-02 | 1.47E-02 | 1.98E-03 | 2.34E-01 | 3.40E-01 | 9.85E-01 |
| G_oc-temp_med-Parahip | 4.22E-06 | 6.23E-02 | 7.29E-03 | 6.41E-03 | -2.77E-06 | 4.38E-02 | -6.00E-02 | 5.15E-02 | 6.79E-01 | 4.17E-01 | 7.08E-01 | 3.38E-01 |
| G_occipital_middle | 2.91E-05 | 6.88E-11 | 3.65E-02 | 6.09E-10 | -1.32E-05 | 1.47E-07 | -2.62E-02 | 1.84E-10 | 1.44E-06 | 5.14E-07 | 7.42E-05 | 3.38E-01 |
| G_occipital_sup | 9.88E-06 | 3.87E-01 | 1.32E-02 | 5.56E-01 | -5.79E-06 | 9.85E-02 | -6.21E-02 | 5.65E-02 | 5.93E-04 | 1.19E-04 | 3.35E-02 | 8.87E-01 |
| G_orbital | -4.28E-07 | 6.79E-01 | 6.60E-04 | 4.52E-02 | -1.61E-06 | 5.49E-03 | -3.04E-02 | 9.03E-03 | 5.27E-01 | 1.86E-01 | 3.69E-01 | 9.85E-01 |
| G_pariet_inf-Angular | 1.88E-05 | 2.80E-08 | 2.11E-02 | 6.10E-07 | -5.59E-06 | 1.73E-02 | -5.05E-02 | 6.66E-01 | 1.44E-06 | 1.10E-07 | 1.24E-02 | 4.50E-01 |
| G_pariet_inf-Supramar | 5.70E-06 | 1.20E-01 | 7.05E-03 | 3.18E-02 | -2.80E-06 | 2.58E-01 | -2.27E-02 | 7.66E-01 | 2.11E-01 | 5.23E-03 | 4.62E-01 | 4.50E-01 |
| G_parietal_sup | -1.23E-05 | 9.49E-02 | -3.50E-03 | 6.71E-02 | -9.03E-06 | 2.44E-01 | -8.80E-02 | 1.43E-01 | 3.48E-01 | 8.47E-01 | 2.05E-01 | 7.26E-01 |
| G_postcentral | -1.53E-05 | 3.28E-01 | -3.46E-03 | 1.30E-01 | -1.31E-05 | 5.16E-01 | 1.70E-02 | 5.97E-02 | 2.39E-01 | 8.41E-01 | 2.99E-02 | 3.38E-01 |
| G_precentral | -1.50E-05 | 2.93E-02 | -2.52E-03 | 1.45E-01 | -1.33E-05 | 3.20E-02 | -8.12E-02 | 3.25E-01 | 2.59E-01 | 7.94E-01 | 7.93E-02 | 9.43E-01 |
| G_precuneus | -5.61E-07 | 4.95E-01 | 1.59E-03 | 8.11E-01 | -1.44E-06 | 4.84E-01 | -8.17E-02 | 2.18E-01 | 8.56E-01 | 3.97E-01 | 4.57E-01 | 9.85E-01 |
| G_rectus | -7.54E-07 | 8.88E-01 | 4.38E-06 | 4.58E-01 | -7.13E-07 | 1.34E-01 | -8.53E-02 | 1.82E-05 | 8.37E-01 | 4.03E-01 | 3.69E-01 | 5.10E-01 |
| G_subcallosal | -1.78E-05 | 8.32E-14 | -2.47E-02 | 2.89E-13 | 7.69E-06 | 8.80E-04 | 1.10E-01 | 4.63E-01 | 2.92E-01 | 6.47E-02 | 2.12E-01 | 5.71E-01 |
| G_temp_sup-G_T_transv | 4.46E-06 | 5.24E-03 | 3.60E-05 | 3.21E-01 | 3.04E-06 | 3.53E-01 | -1.98E-01 | 6.41E-01 | 9.29E-01 | 3.44E-01 | 1.91E-02 | 9.85E-01 |
| G_temp_sup-Lateral | 4.38E-06 | 1.19E-01 | 2.78E-03 | 6.21E-01 | 2.21E-06 | 9.45E-02 | -4.06E-02 | 4.16E-01 | 3.53E-01 | 3.06E-01 | 6.72E-01 | 7.01E-01 |
| G_temp_sup-Plan_polar | 6.82E-06 | 1.31E-04 | 9.12E-03 | 3.13E-04 | -4.30E-06 | 2.99E-05 | 1.58E-01 | 2.00E-02 | 9.40E-01 | 9.25E-01 | 5.30E-01 | 4.62E-01 |
| G_temp_sup-Plan_tempo | 4.34E-06 | 1.34E-03 | 3.77E-03 | 2.30E-02 | -6.78E-07 | 7.52E-02 | 2.86E-02 | 7.66E-01 | 9.80E-01 | 7.56E-01 | 4.62E-01 | 3.38E-01 |
| G_temporal_inf | 9.35E-06 | 4.02E-03 | 1.05E-02 | 1.65E-02 | -3.55E-06 | 1.73E-02 | -1.17E-02 | 2.18E-01 | 2.39E-01 | 2.78E-01 | 7.08E-01 | 9.85E-01 |
| G_temporal_middle | 8.15E-06 | 2.42E-07 | 6.83E-03 | 2.74E-04 | 8.67E-07 | 7.38E-01 | -9.36E-02 | 3.01E-01 | 3.18E-01 | 6.28E-01 | 7.83E-01 | 8.94E-01 |

| | | | | | | | | | | | | |
|---|---|---|---|---|---|---|---|---|---|---|---|---|
| Lat_Fis-ant-Horizont | -3.66E-07 | 7.92E-01 | 3.65E-03 | 9.25E-01 | 2.97E-07 | 9.08E-01 | -8.86E-03 | 5.17E-04 | 8.92E-01 | 5.58E-01 | 9.87E-01 | 4.50E-01 |
| Lat_Fis-ant-Vertical | 1.13E-06 | 5.63E-01 | 1.42E-03 | 7.87E-01 | -4.59E-07 | 8.40E-01 | -5.42E-02 | 9.03E-01 | 9.67E-01 | 9.05E-01 | 8.48E-01 | 6.54E-01 |
| Lat_Fis-post | 7.38E-06 | 3.74E-06 | 9.43E-03 | 5.18E-04 | -3.12E-06 | 9.49E-05 | -7.41E-02 | 3.90E-01 | 1.25E-01 | 9.84E-02 | 2.12E-01 | 1.69E-01 |
| Pole_occipital | 2.44E-05 | 1.71E-13 | 3.22E-02 | 5.96E-17 | -1.15E-05 | 1.57E-04 | 7.82E-02 | 1.18E-01 | 4.45E-03 | 2.35E-06 | 7.42E-05 | 4.50E-01 |
| Pole_temporal | 2.24E-06 | 8.81E-01 | 3.78E-03 | 6.75E-01 | -1.20E-06 | 8.37E-01 | -7.90E-02 | 2.25E-02 | 4.90E-01 | 5.16E-01 | 5.62E-01 | 8.87E-01 |
| S_calcarine | -7.04E-07 | 1.30E-01 | -4.98E-04 | 1.56E-01 | 5.04E-07 | 9.79E-02 | -2.45E-03 | 8.46E-06 | 3.14E-01 | 2.56E-01 | 3.21E-01 | 8.05E-01 |
| S_central | 5.87E-06 | 8.21E-03 | 6.74E-03 | 5.76E-02 | -2.80E-07 | 9.89E-01 | 1.18E-02 | 9.55E-02 | 2.10E-01 | 2.75E-01 | 7.83E-01 | 6.51E-01 |
| S_cingul-Marginalis | -2.01E-06 | 1.10E-03 | -2.62E-03 | 4.89E-03 | 7.33E-07 | 3.20E-02 | 5.58E-02 | 3.01E-01 | 2.25E-01 | 2.34E-01 | 3.31E-01 | 8.05E-01 |
| S_circular_insula_ant | -7.16E-06 | 7.45E-05 | -1.28E-02 | 2.34E-05 | 3.68E-06 | 4.79E-04 | 1.04E-01 | 2.02E-01 | 7.50E-01 | 8.43E-01 | 7.83E-01 | 5.71E-01 |
| S_circular_insula_inf | 6.02E-06 | 9.99E-03 | 8.70E-03 | 3.33E-02 | -2.90E-06 | 3.66E-02 | 3.69E-02 | 4.73E-05 | 3.27E-01 | 2.08E-01 | 2.55E-01 | 8.87E-01 |
| S_circular_insula_sup | -3.04E-06 | 1.12E-04 | -2.67E-03 | 4.89E-03 | 1.54E-06 | 1.12E-04 | 2.54E-02 | 1.89E-05 | 6.79E-01 | 3.35E-01 | 5.62E-01 | 5.10E-01 |
| S_collat_transv_ant | 5.10E-06 | 8.88E-01 | 4.78E-03 | 8.11E-01 | -2.21E-06 | 9.89E-01 | -6.49E-02 | 4.18E-01 | 2.11E-01 | 2.34E-01 | 2.12E-01 | 8.87E-01 |
| S_collat_transv_post | 2.26E-05 | 6.87E-11 | 3.12E-02 | 4.09E-10 | -1.15E-05 | 1.24E-10 | -3.38E-03 | 1.14E-03 | 8.38E-04 | 8.05E-03 | 6.33E-04 | 4.62E-01 |
| S_front_inf | -5.67E-07 | 2.43E-01 | 2.44E-03 | 8.58E-01 | -3.61E-06 | 6.46E-02 | -4.80E-02 | 1.82E-05 | 4.90E-01 | 9.84E-02 | 1.24E-02 | 4.62E-01 |
| S_front_middle | 1.16E-06 | 8.88E-01 | 1.34E-03 | 8.11E-01 | -2.75E-06 | 2.14E-01 | -1.77E-02 | 7.32E-01 | 7.34E-01 | 7.56E-01 | 4.62E-01 | 4.50E-01 |
| S_front_sup | 5.68E-07 | 8.81E-01 | 5.40E-04 | 9.01E-01 | -1.22E-06 | 9.89E-01 | -7.28E-02 | 1.10E-02 | 8.91E-01 | 8.47E-01 | 3.48E-01 | 4.50E-01 |
| S_interm_prim-Jensen | 6.17E-06 | 8.88E-01 | 1.02E-02 | 7.75E-01 | -3.02E-06 | 9.01E-01 | -4.53E-02 | 1.13E-07 | 2.14E-02 | 8.05E-03 | 3.35E-02 | 6.91E-01 |
| S_intrapariet_and_P_trans | 5.63E-06 | 6.24E-01 | 1.24E-02 | 1.66E-01 | -8.65E-06 | 3.95E-04 | 1.44E-02 | 6.38E-02 | 2.32E-02 | 1.17E-05 | 1.02E-07 | 4.50E-01 |
| S_oc-temp_lat | 2.00E-05 | 4.96E-10 | 2.70E-02 | 3.99E-09 | -1.06E-05 | 7.37E-10 | -9.63E-03 | 1.34E-02 | 1.94E-01 | 2.34E-01 | 2.05E-01 | 9.85E-01 |
| S_oc-temp_med_and_Lingual | 9.58E-06 | 1.71E-13 | 1.33E-02 | 1.69E-13 | -4.72E-06 | 1.25E-10 | 2.54E-02 | 6.66E-01 | 3.48E-01 | 4.17E-01 | 1.88E-01 | 6.20E-01 |
| S_oc_middle_and_Lunatus | 3.74E-05 | 1.14E-26 | 4.79E-02 | 1.59E-22 | -1.81E-05 | 3.14E-24 | 3.40E-02 | 1.94E-08 | 1.60E-04 | 1.02E-03 | 7.42E-05 | 3.38E-01 |
| S_oc_sup_and_transversal | 1.72E-05 | 5.27E-06 | 2.37E-02 | 1.35E-05 | -8.32E-06 | 8.73E-05 | -1.13E-03 | 5.15E-02 | 3.54E-05 | 1.03E-04 | 6.51E-05 | 6.20E-01 |
| S_occipital_ant | 2.63E-05 | 5.89E-10 | 3.45E-02 | 9.77E-09 | -1.26E-05 | 1.41E-07 | -5.88E-03 | 1.16E-01 | 4.45E-03 | 2.67E-03 | 4.84E-03 | 5.71E-01 |
| S_orbital-H_Shaped | 2.45E-06 | 3.39E-02 | 3.20E-03 | 1.32E-02 | -8.86E-07 | 3.61E-02 | -1.68E-02 | 7.66E-01 | 4.57E-01 | 2.56E-01 | 2.79E-01 | 7.01E-01 |
| S_orbital_lateral | 4.83E-06 | 2.93E-02 | 7.93E-03 | 2.82E-02 | -1.81E-06 | 4.79E-02 | -5.17E-02 | 4.18E-01 | 5.63E-01 | 6.19E-01 | 3.69E-01 | 5.10E-01 |
| S_orbital_med_olfact | -4.58E-06 | 6.24E-01 | -5.74E-03 | 8.11E-01 | 1.13E-06 | 9.08E-01 | 7.49E-02 | 4.09E-01 | 7.50E-01 | 6.53E-01 | 5.96E-01 | 9.43E-01 |
| S_parieto_occipital | 1.38E-06 | 8.88E-01 | 4.47E-03 | 3.21E-01 | -3.73E-07 | 8.05E-01 | -6.45E-02 | 3.92E-01 | 3.46E-01 | 2.08E-01 | 3.69E-01 | 9.85E-01 |
| S_pericallosal | -1.03E-05 | 2.07E-04 | -1.47E-02 | 5.28E-07 | 4.78E-06 | 1.13E-06 | 7.65E-02 | 1.51E-03 | 3.48E-01 | 3.44E-01 | 2.79E-01 | 5.71E-01 |
| S_postcentral | 1.32E-06 | 5.04E-01 | 3.11E-04 | 1.99E-02 | 7.09E-07 | 6.24E-02 | -2.56E-02 | 1.84E-10 | 1.05E-01 | 3.98E-03 | 2.18E-01 | 8.05E-01 |
| S_precentral-inf-part | 2.56E-07 | 1.17E-01 | 8.36E-04 | 6.71E-02 | -1.70E-06 | 1.39E-01 | -5.68E-02 | 6.46E-05 | 1.88E-01 | 2.25E-01 | 8.48E-01 | 4.50E-01 |
| S_precentral-sup-part | -5.77E-07 | 8.06E-01 | -3.77E-03 | 4.65E-02 | 1.25E-06 | 1.23E-01 | -6.77E-02 | 5.41E-03 | 9.80E-01 | 7.56E-01 | 5.62E-01 | 9.70E-01 |
| S_suborbital | -2.19E-06 | 3.87E-01 | -4.16E-03 | 6.21E-01 | 1.74E-06 | 7.51E-01 | -8.63E-02 | 3.92E-01 | 8.05E-02 | 8.80E-02 | 7.87E-02 | 9.85E-01 |
| S_subparietal | -6.31E-07 | 2.09E-02 | -7.52E-04 | 1.67E-02 | 2.35E-07 | 2.38E-02 | 6.88E-02 | 5.17E-04 | 1.19E-01 | 6.27E-02 | 7.87E-02 | 4.50E-01 |
| S_temporal_inf | 8.50E-06 | 1.08E-01 | 9.43E-03 | 3.90E-01 | -4.31E-06 | 1.64E-01 | 1.90E-02 | 7.43E-01 | 4.08E-01 | 3.97E-01 | 3.40E-01 | 5.52E-01 |
| S_temporal_sup | 8.09E-06 | 6.23E-04 | 9.65E-03 | 1.91E-02 | -3.29E-06 | 2.99E-02 | -1.53E-02 | 1.72E-03 | 1.01E-04 | 1.19E-04 | 5.60E-04 | 6.54E-01 |
| S_temporal_transverse | 4.83E-06 | 2.09E-02 | 6.33E-03 | 4.65E-02 | -6.90E-07 | 6.04E-01 | -4.33E-02 | 2.20E-02 | 5.85E-01 | 6.54E-01 | 8.48E-01 | 5.95E-01 |

**Summary of with lateralization in diffusion measures and cortical thickness (left) and significance level of the AGE_GROUP × SIDE interaction (right) in the combined group.**
Displayed is the effect of *SIDE* with according adjusted P-value (false-discovery-rate). For the interaction the adjusted P-value (false-discovery-rate) is given. AD: axial diffusivity, RD: radial diffusivity, FA: fractional anisotropy, CT: cortical thickness.

**Table S2.** Result overview, part II.

| Region | AD younger adults | | AD older adults | | FA younger adults | | FA older adults | | RD younger adults | | RD older adults | | CT younger adults | | CT older adults | |
|---|---|---|---|---|---|---|---|---|---|---|---|---|---|---|---|---|
| | coefficient | $P_{FDR}$ | coefficient | $P_{FDR}$ | coefficient | $P_{FDR}$ | coefficient | $P_{FDR}$ | coefficient | $P_{FDR}$ | coefficient | $P_{FDR}$ | coefficient | $P_{FDR}$ | coefficient | $P_{FDR}$ |
| G&S_cingul-Ant | 2.20E-07 | 9.07E-01 | -1.04E-06 | 4.25E-01 | 6.59E-04 | 7.93E-01 | -2.22E-03 | 2.68E-01 | -3.15E-07 | 7.42E-01 | 4.96E-07 | 4.56E-01 | 3.62E-02 | 1.28E-02 | -2.17E-02 | 7.19E-04 |
| G&S_cingul-Mid-Ant | 3.49E-06 | 1.49E-02 | 1.31E-06 | 4.27E-01 | 4.91E-03 | 3.79E-02 | 1.31E-03 | 6.10E-01 | -1.94E-06 | 4.20E-02 | -1.59E-06 | 4.02E-02 | -1.68E-02 | 4.29E-01 | 3.23E-03 | 4.45E-01 |
| G&S_cingul-Mid-Post | -1.06E-06 | 6.56E-01 | -1.14E-06 | 4.74E-01 | -1.66E-03 | 6.34E-01 | 1.47E-03 | 5.90E-01 | 1.78E-07 | 9.37E-01 | -9.43E-07 | 3.39E-01 | 3.09E-02 | 9.34E-01 | 2.75E-02 | 1.31E-01 |
| G&S_frontomargin | -2.06E-06 | 5.21E-01 | -2.99E-06 | 6.31E-01 | -1.18E-03 | 7.75E-01 | -1.72E-03 | 6.52E-01 | -2.81E-06 | 2.58E-01 | -1.39E-06 | 7.45E-01 | -5.88E-03 | 7.71E-01 | -5.76E-02 | 5.63E-01 |
| G&S_occipital_inf | 1.14E-05 | 1.40E-02 | 3.68E-05 | 5.14E-14 | 1.66E-02 | 5.36E-03 | 5.33E-02 | 8.97E-26 | -6.13E-06 | 1.25E-02 | -2.37E-05 | 4.50E-12 | -3.65E-02 | 4.28E-06 | 5.84E-02 | 6.51E-07 |
| G&S_paracentral | -1.63E-06 | 7.17E-01 | -7.97E-06 | 1.03E-01 | -7.42E-03 | 5.91E-03 | -2.83E-03 | 3.73E-01 | 5.08E-06 | 7.08E-02 | -5.56E-06 | 2.18E-01 | -3.12E-02 | 9.03E-01 | 5.37E-02 | 4.21E-05 |
| G&S_subcentral | -2.29E-07 | 9.07E-01 | 2.61E-06 | 2.54E-02 | -9.38E-04 | 6.61E-01 | 1.58E-03 | 4.06E-01 | 5.14E-07 | 6.99E-01 | 1.67E-06 | 1.12E-01 | -2.10E-02 | 1.69E-03 | -1.03E-02 | 2.24E-01 |
| G&S_transv_frontopol | 3.83E-06 | 4.72E-01 | 5.19E-06 | 7.04E-01 | 1.58E-03 | 5.92E-01 | -7.14E-03 | 1.54E-01 | 2.38E-06 | 6.34E-01 | 1.09E-05 | 2.10E-01 | -2.79E-02 | 3.77E-02 | -6.75E-02 | 5.97E-01 |
| G_cingul-Post-dorsal | -3.58E-06 | 6.05E-03 | -1.74E-06 | 1.74E-01 | -6.36E-03 | 1.91E-03 | -1.94E-03 | 3.56E-01 | 1.86E-06 | 5.79E-03 | 7.32E-07 | 2.95E-01 | 1.71E-02 | 9.95E-03 | 1.43E-02 | 5.12E-01 |
| G_cingul-Post-ventral | -1.38E-05 | 3.66E-06 | -6.38E-06 | 1.39E-02 | -2.03E-02 | 5.90E-07 | -7.21E-03 | 8.90E-02 | 5.46E-06 | 9.97E-05 | 2.09E-06 | 1.70E-01 | -3.32E-02 | 7.61E-01 | 1.13E-02 | 4.80E-02 |
| G_cuneus | -4.48E-06 | 7.11E-02 | 3.67E-06 | 2.03E-01 | -5.09E-03 | 1.46E-01 | 7.47E-03 | 8.10E-03 | 2.24E-06 | 5.11E-02 | -3.10E-07 | 8.24E-01 | 1.10E-02 | 3.90E-01 | 4.60E-02 | 2.09E-01 |
| G_front_inf-Opercular | 5.21E-07 | 8.70E-01 | 2.47E-06 | 6.31E-01 | 3.24E-04 | 9.02E-01 | -1.41E-03 | 5.37E-01 | -5.64E-07 | 6.86E-01 | 3.04E-06 | 4.60E-01 | 2.86E-02 | 7.19E-02 | 4.94E-02 | 5.77E-01 |
| G_front_inf-Orbital | 2.42E-07 | 9.81E-01 | -3.33E-06 | 6.31E-01 | 7.61E-04 | 9.02E-01 | -1.38E-03 | 6.99E-01 | 8.95E-08 | 9.64E-01 | -4.37E-07 | 9.07E-01 | -4.44E-02 | 9.34E-01 | 2.17E-02 | 5.12E-01 |
| G_front_inf-Triangul | -2.89E-06 | 1.13E-01 | -2.23E-06 | 6.46E-01 | -6.26E-03 | 3.13E-03 | -2.57E-03 | 4.44E-01 | 1.57E-06 | 3.13E-01 | -1.18E-06 | 7.40E-01 | 1.10E-01 | 2.30E-01 | 4.47E-04 | 1.03E-01 |
| G_front_middle | 4.35E-07 | 8.85E-01 | -1.72E-06 | 6.31E-01 | 4.51E-04 | 8.16E-01 | -7.62E-04 | 7.30E-01 | -4.14E-07 | 8.09E-01 | -1.40E-06 | 6.17E-01 | 6.81E-03 | 4.08E-02 | -6.69E-03 | 9.61E-01 |
| G_front_sup | 3.47E-07 | 8.85E-01 | -7.95E-06 | 1.22E-02 | 1.46E-04 | 9.06E-01 | -3.43E-03 | 7.53E-03 | 8.76E-08 | 9.64E-01 | -4.91E-06 | 6.76E-02 | 3.28E-02 | 1.83E-01 | -2.82E-02 | 5.76E-01 |
| G_Ins_lg&S_cent_ins | -8.02E-06 | 2.30E-04 | -5.71E-06 | 9.55E-02 | -1.07E-02 | 1.18E-03 | -6.43E-03 | 1.73E-01 | 3.98E-06 | 3.55E-04 | 3.16E-06 | 7.58E-02 | -2.90E-02 | 1.00E-01 | -2.58E-02 | 8.87E-06 |
| G_insular_short | -6.18E-06 | 2.20E-03 | -4.63E-06 | 7.20E-02 | -7.72E-03 | 7.48E-03 | -6.77E-03 | 5.59E-02 | 2.92E-06 | 5.04E-03 | 1.93E-06 | 1.69E-01 | -5.88E-02 | 1.52E-04 | 7.60E-02 | 2.12E-02 |
| G_occipital_middle | 3.02E-06 | 5.11E-01 | 2.94E-05 | 2.60E-15 | 4.09E-03 | 4.28E-01 | 3.62E-02 | 4.65E-13 | -3.72E-06 | 5.11E-02 | -1.25E-05 | 3.99E-09 | 1.50E-02 | 5.43E-18 | 2.47E-02 | 4.21E-05 |
| G_occipital_sup | -7.05E-06 | 3.36E-03 | 1.04E-05 | 3.41E-03 | -1.14E-02 | 2.02E-05 | 1.42E-02 | 5.35E-03 | 1.76E-06 | 2.74E-01 | -6.07E-06 | 1.81E-02 | -3.82E-02 | 1.56E-02 | 7.98E-02 | 1.15E-01 |
| G_oc-temp_lat-fusifor | 3.41E-06 | 9.31E-03 | 9.21E-06 | 9.46E-07 | 5.76E-03 | 1.16E-03 | 1.39E-02 | 1.01E-07 | -3.42E-06 | 1.05E-04 | -6.45E-06 | 4.73E-07 | 4.65E-02 | 3.99E-01 | 1.45E-02 | 4.12E-03 |
| G_oc-temp_med-Lingual | 1.50E-06 | 2.17E-01 | 9.67E-06 | 9.31E-07 | 1.49E-03 | 4.01E-01 | 5.21E-03 | 2.17E-03 | 5.11E-07 | 6.34E-01 | 3.07E-06 | 8.36E-02 | 9.50E-02 | 8.45E-02 | -1.46E-03 | 3.35E-02 |
| G_oc-temp_med-Parahip | 2.15E-06 | 4.79E-01 | 3.59E-06 | 6.13E-02 | 3.53E-03 | 2.96E-01 | 6.28E-03 | 1.64E-03 | -1.76E-06 | 2.34E-01 | -2.52E-06 | 1.34E-01 | -1.42E-01 | 9.96E-01 | -8.22E-02 | 3.77E-03 |
| G_orbital | 2.47E-06 | 1.72E-01 | -6.18E-08 | 9.73E-01 | 5.53E-03 | 1.94E-03 | 9.95E-04 | 6.19E-01 | -3.62E-06 | 1.13E-04 | -1.88E-06 | 2.31E-01 | -2.35E-02 | 1.06E-01 | -3.15E-02 | 8.79E-03 |
| G_pariet_inf-Angular | -7.09E-09 | 9.96E-01 | 1.82E-05 | 1.34E-14 | -2.22E-03 | 3.73E-01 | 2.05E-02 | 6.83E-13 | 4.24E-07 | 7.42E-01 | -5.67E-06 | 2.04E-03 | -4.88E-02 | 3.24E-01 | 5.53E-02 | 6.09E-02 |
| G_pariet_inf-Supramar | 4.99E-07 | 8.12E-01 | 5.94E-06 | 6.13E-02 | -1.08E-03 | 5.92E-01 | 7.23E-03 | 1.12E-03 | -5.24E-08 | 9.64E-01 | -2.66E-06 | 3.12E-01 | 1.99E-02 | 1.06E-01 | 7.95E-02 | 3.57E-01 |
| G_parietal_sup | -2.50E-06 | 3.68E-01 | -1.33E-05 | 9.50E-02 | -4.22E-03 | 3.49E-02 | -4.13E-03 | 3.04E-01 | 8.15E-07 | 8.09E-01 | -9.31E-06 | 1.56E-01 | -3.26E-02 | 4.26E-01 | 8.64E-02 | 1.13E-03 |
| G_postcentral | -6.13E-07 | 9.07E-01 | -1.52E-05 | 2.08E-01 | -5.72E-03 | 1.20E-02 | -3.40E-03 | 5.37E-01 | 4.45E-06 | 1.45E-01 | -1.31E-05 | 1.02E-01 | -2.07E-02 | 1.52E-04 | 2.88E-02 | 8.22E-01 |
| G_precentral | -2.50E-06 | 1.79E-01 | -1.53E-05 | 6.13E-02 | -3.76E-03 | 8.12E-02 | -2.60E-03 | 5.37E-01 | -2.75E-07 | 9.33E-01 | -1.36E-05 | 2.73E-02 | 2.71E-02 | 4.38E-01 | 2.84E-03 | 4.45E-01 |
| G_precuneus | -1.29E-06 | 4.37E-01 | -6.35E-07 | 7.08E-01 | -9.46E-04 | 5.87E-01 | 1.54E-03 | 4.85E-01 | 1.65E-07 | 9.37E-01 | -1.52E-06 | 2.55E-01 | 3.30E-02 | 2.30E-01 | 7.76E-02 | 3.05E-01 |
| G_rectus | -1.53E-07 | 9.81E-01 | -2.64E-07 | 9.25E-01 | 2.61E-03 | 3.73E-01 | 1.18E-03 | 5.89E-01 | -3.40E-06 | 1.72E-02 | -1.02E-06 | 6.17E-01 | -6.41E-03 | 2.00E-04 | -2.13E-03 | 1.46E-02 |
| G_subcallosal | -1.19E-05 | 7.08E-06 | -1.78E-05 | 6.16E-12 | -1.26E-02 | 7.14E-05 | -2.47E-02 | 1.66E-13 | 2.60E-06 | 3.20E-01 | 7.73E-06 | 3.44E-07 | -1.71E-02 | 9.03E-01 | -1.90E-02 | 2.11E-01 |
| G_temp_sup-G_T_transv | 3.72E-06 | 4.07E-02 | 4.60E-06 | 4.33E-02 | 5.12E-03 | 8.37E-02 | 1.77E-04 | 9.48E-01 | -1.49E-06 | 1.63E-01 | 3.03E-06 | 1.77E-02 | -1.48E-01 | 8.20E-01 | 5.37E-02 | 5.98E-01 |
| G_temp_sup-Lateral | 6.54E-07 | 8.12E-01 | 4.38E-06 | 4.12E-02 | -1.05E-03 | 5.83E-01 | 2.83E-03 | 2.12E-01 | 1.19E-06 | 3.19E-01 | 2.00E-06 | 1.88E-01 | -1.34E-02 | 9.08E-01 | -2.36E-02 | 2.64E-01 |
| G_temp_sup-Plan_polar | 6.27E-06 | 1.97E-02 | 6.76E-06 | 1.61E-03 | 9.24E-03 | 9.59E-03 | 8.88E-03 | 1.65E-03 | -2.94E-06 | 3.49E-02 | -4.24E-06 | 1.81E-05 | 1.00E-01 | 3.54E-03 | 1.88E-02 | 4.73E-01 |
| G_temp_sup-Plan_tempo | 4.39E-06 | 2.41E-02 | 4.40E-06 | 3.59E-03 | 5.61E-03 | 3.79E-02 | 3.67E-03 | 1.47E-01 | -2.02E-06 | 4.08E-02 | -6.09E-07 | 5.59E-01 | 8.27E-03 | 2.29E-01 | 9.73E-02 | 1.86E-01 |
| G_temporal_inf | 3.16E-06 | 3.12E-01 | 9.40E-06 | 5.05E-04 | 3.46E-03 | 3.73E-01 | 1.04E-02 | 1.99E-03 | -2.58E-06 | 5.11E-02 | -3.39E-06 | 4.46E-02 | -6.76E-02 | 1.86E-01 | -1.05E-02 | 2.11E-01 |
| G_temporal_middle | 4.59E-06 | 3.17E-03 | 8.20E-06 | 1.51E-06 | 4.65E-03 | 1.20E-02 | 6.91E-03 | 7.21E-04 | 1.57E-07 | 9.37E-01 | 7.68E-07 | 6.45E-01 | 2.51E-02 | 5.47E-01 | -2.11E-02 | 2.03E-01 |
| Lat_Fis-ant-Horizont | -2.26E-06 | 7.58E-01 | -6.08E-07 | 8.86E-01 | -3.28E-03 | 7.67E-01 | 2.96E-03 | 6.10E-01 | 6.26E-07 | 9.33E-01 | 5.10E-07 | 7.80E-01 | 5.37E-02 | 1.52E-04 | -6.11E-02 | 2.06E-01 |
| Lat_Fis-ant-Vertical | 1.34E-06 | 7.37E-01 | 1.06E-06 | 6.63E-01 | 4.05E-04 | 9.19E-01 | 1.31E-03 | 6.74E-01 | -5.74E-08 | 9.64E-01 | -4.36E-07 | 7.20E-01 | 5.63E-02 | 6.02E-01 | 7.20E-02 | 5.77E-01 |

| | | | | | | | | | | | | | | | | |
|---|---|---|---|---|---|---|---|---|---|---|---|---|---|---|---|---|
| Lat_Fis-post | 2.94E-06 | 3.18E-02 | 7.77E-06 | 3.29E-06 | 2.58E-03 | 2.63E-01 | 1.01E-02 | 2.37E-05 | -1.28E-06 | 6.97E-02 | -3.30E-06 | 6.16E-05 | 5.67E-03 | 2.91E-01 | 2.47E-02 | 8.27E-04 |
| Pole_occipital | 5.50E-06 | 7.11E-02 | 2.47E-05 | 2.50E-19 | 4.36E-03 | 2.02E-01 | 3.24E-02 | 8.97E-26 | 5.27E-07 | 9.33E-01 | -1.14E-05 | 8.92E-13 | 5.55E-03 | 1.80E-04 | 6.49E-03 | 9.43E-01 |
| Pole_temporal | -1.69E-06 | 5.11E-01 | 2.27E-06 | 4.41E-01 | -4.36E-04 | 9.06E-01 | 3.16E-03 | 4.44E-01 | 5.75E-08 | 9.64E-01 | -7.96E-07 | 6.80E-01 | -1.92E-01 | 1.37E-01 | 5.47E-02 | 4.80E-02 |
| S_calcarine | -5.45E-06 | 5.27E-03 | 3.54E-07 | 9.05E-01 | -8.12E-03 | 5.50E-03 | 1.53E-03 | 6.74E-01 | 2.75E-06 | 5.79E-03 | -4.02E-08 | 9.70E-01 | 2.60E-02 | 1.40E-02 | 1.75E-02 | 3.79E-07 |
| S_central | 1.61E-06 | 3.49E-01 | 5.82E-06 | 1.04E-02 | 1.79E-03 | 4.95E-01 | 6.64E-03 | 4.16E-02 | 1.95E-07 | 9.33E-01 | -2.73E-07 | 8.28E-01 | 2.50E-02 | 8.01E-02 | 6.91E-02 | 5.41E-01 |
| S_cingul-Marginalis | -5.67E-06 | 1.89E-03 | -2.00E-06 | 2.41E-01 | -8.19E-03 | 3.23E-03 | -2.56E-03 | 3.56E-01 | 2.52E-06 | 4.08E-02 | 7.25E-07 | 4.60E-01 | 2.61E-02 | 2.49E-01 | 1.08E-01 | 5.77E-01 |
| S_circular_insula_ant | -8.54E-06 | 2.41E-02 | -7.21E-06 | 2.28E-03 | -1.35E-02 | 2.26E-02 | -1.29E-02 | 1.39E-04 | 4.08E-06 | 5.94E-02 | 3.72E-06 | 1.19E-03 | 4.83E-02 | 1.17E-01 | 4.04E-02 | 7.49E-01 |
| S_circular_insula_inf | 2.90E-06 | 2.30E-01 | 5.68E-06 | 3.54E-03 | 2.85E-03 | 4.28E-01 | 8.07E-03 | 8.10E-03 | -1.02E-06 | 4.50E-01 | -2.73E-06 | 1.30E-02 | 2.72E-03 | 3.75E-03 | -4.75E-03 | 1.13E-03 |
| S_circular_insula_sup | -4.12E-06 | 9.46E-04 | -3.04E-06 | 2.75E-02 | -6.19E-03 | 1.86E-03 | -2.51E-03 | 3.04E-01 | 2.17E-06 | 1.22E-03 | 1.54E-06 | 2.85E-02 | 7.11E-02 | 3.28E-02 | -2.29E-03 | 8.67E-10 |
| S_collat_transv_ant | -4.53E-06 | 2.66E-01 | 5.11E-06 | 3.09E-01 | -8.00E-03 | 1.46E-01 | 4.74E-03 | 5.37E-01 | 2.53E-06 | 1.63E-01 | -2.14E-06 | 4.14E-01 | 4.75E-02 | 7.61E-01 | -6.32E-02 | 4.71E-01 |
| S_collat_transv_post | 6.32E-06 | 1.69E-02 | 2.19E-05 | 2.61E-11 | 1.12E-02 | 3.49E-03 | 3.06E-02 | 1.36E-09 | -3.16E-06 | 1.83E-02 | -1.12E-05 | 2.86E-11 | -8.09E-03 | 1.24E-01 | -4.37E-03 | 2.26E-03 |
| S_front_inf | -2.70E-06 | 3.20E-02 | -7.15E-07 | 7.08E-01 | -3.05E-03 | 1.55E-01 | 2.41E-03 | 3.04E-01 | 3.40E-07 | 8.29E-01 | -3.59E-06 | 4.99E-03 | 7.55E-02 | 5.02E-06 | 1.24E-03 | 1.14E-02 |
| S_front_middle | -2.13E-07 | 9.71E-01 | 1.30E-06 | 7.04E-01 | 6.38E-04 | 8.18E-01 | 1.39E-03 | 6.19E-01 | -6.07E-07 | 7.13E-01 | -2.75E-06 | 1.88E-01 | 6.36E-02 | 1.83E-01 | -1.01E-02 | 5.41E-01 |
| S_front_sup | 1.46E-06 | 2.15E-01 | 4.67E-07 | 8.53E-01 | 2.02E-03 | 2.34E-01 | 4.11E-04 | 8.23E-01 | 5.78E-07 | 5.77E-01 | -1.12E-06 | 5.75E-01 | 4.19E-02 | 2.44E-04 | 3.86E-02 | 2.79E-01 |
| S_interm_prim-Jensen | -7.14E-06 | 1.87E-02 | 7.16E-06 | 9.14E-02 | -9.77E-03 | 1.39E-02 | 1.15E-02 | 5.44E-02 | 3.58E-06 | 1.96E-02 | -3.89E-06 | 8.07E-02 | 1.17E-01 | 1.52E-04 | 1.70E-01 | 7.19E-04 |
| S_intrapariet&P_trans | -3.61E-06 | 5.84E-03 | 5.15E-06 | 1.17E-01 | -6.63E-03 | 1.91E-03 | 1.14E-02 | 9.75E-04 | 2.01E-06 | 9.68E-03 | -8.40E-06 | 4.73E-07 | 2.51E-02 | 7.61E-01 | 1.08E-01 | 1.13E-03 |
| S_oc_middle&Lunatus | 1.10E-05 | 3.29E-03 | 3.96E-05 | 4.56E-28 | 1.42E-02 | 7.72E-03 | 5.10E-02 | 1.73E-24 | -4.63E-06 | 1.77E-02 | -1.91E-05 | 1.34E-28 | 4.57E-02 | 3.36E-08 | 5.48E-02 | 1.13E-03 |
| S_oc_sup&transversal | -4.80E-07 | 8.98E-01 | 1.72E-05 | 3.30E-08 | -4.18E-04 | 9.09E-01 | 2.38E-02 | 5.75E-08 | 6.43E-07 | 6.99E-01 | -8.32E-06 | 3.35E-07 | 2.34E-02 | 2.95E-02 | 1.34E-01 | 3.01E-01 |
| S_occipital_ant | 7.79E-06 | 4.07E-02 | 2.63E-05 | 9.57E-13 | 8.57E-03 | 8.73E-02 | 3.44E-02 | 6.04E-12 | -2.87E-06 | 1.37E-01 | -1.26E-05 | 5.90E-11 | -7.69E-03 | 3.28E-02 | 4.99E-02 | 5.92E-01 |
| S_oc-temp_lat | 1.11E-05 | 1.48E-03 | 2.02E-05 | 6.12E-12 | 1.55E-02 | 3.23E-03 | 2.74E-02 | 2.38E-11 | -5.89E-06 | 1.80E-03 | -1.08E-05 | 6.08E-12 | 3.68E-02 | 4.89E-02 | 6.82E-02 | 4.49E-02 |
| S_oc-temp_med&Lingual | 6.95E-06 | 1.60E-05 | 9.56E-06 | 2.39E-13 | 1.04E-02 | 5.90E-07 | 1.33E-02 | 9.32E-12 | -2.67E-06 | 3.96E-03 | -4.70E-06 | 3.24E-13 | 1.23E-02 | 3.20E-01 | 2.98E-02 | 8.09E-01 |
| S_orbital_lateral | 1.01E-05 | 8.89E-02 | 4.71E-06 | 2.12E-01 | 1.43E-02 | 9.19E-02 | 7.76E-03 | 1.66E-01 | -6.07E-06 | 5.38E-02 | -1.77E-06 | 4.14E-01 | 5.11E-02 | 2.29E-01 | -4.93E-02 | 8.22E-01 |
| S_orbital_med_olfact | 2.35E-07 | 9.91E-01 | -5.23E-06 | 2.65E-01 | 3.30E-03 | 8.11E-01 | -6.85E-03 | 3.56E-01 | -2.67E-06 | 6.89E-01 | 1.51E-06 | 5.59E-01 | 2.48E-02 | 5.47E-01 | 6.45E-02 | 5.45E-01 |
| S_orbital-H_Shaped | 6.40E-06 | 1.82E-02 | 2.31E-06 | 4.41E-01 | 1.07E-02 | 1.16E-03 | 2.87E-03 | 5.10E-01 | -3.71E-06 | 7.53E-03 | -7.23E-07 | 6.17E-01 | -6.06E-02 | 8.37E-01 | -2.83E-02 | 4.71E-01 |
| S_parieto_occipital | -1.33E-06 | 3.08E-01 | 1.38E-06 | 4.79E-01 | -1.21E-03 | 5.81E-01 | 4.53E-03 | 1.12E-01 | 8.80E-07 | 1.80E-01 | -3.69E-07 | 7.10E-01 | -1.46E-02 | 4.36E-01 | 6.23E-02 | 5.71E-01 |
| S_pericallosal | -1.69E-05 | 2.78E-05 | -1.04E-05 | 3.54E-03 | -2.42E-02 | 4.67E-05 | -1.48E-02 | 5.35E-03 | 8.61E-06 | 9.98E-05 | 4.84E-06 | 1.66E-02 | -1.12E-03 | 3.54E-03 | 3.48E-02 | 1.01E-01 |
| S_postcentral | -3.41E-06 | 7.02E-02 | 4.08E-07 | 9.01E-01 | -9.37E-03 | 2.40E-04 | -7.87E-04 | 7.81E-01 | 3.74E-06 | 1.43E-02 | 1.11E-06 | 5.75E-01 | 3.23E-02 | 1.14E-11 | 1.04E-01 | 8.67E-10 |
| S_precentral-inf-part | 6.01E-06 | 3.31E-04 | 1.01E-07 | 9.73E-01 | 8.24E-03 | 9.60E-04 | 4.78E-04 | 8.41E-01 | -2.20E-06 | 1.77E-02 | -1.54E-06 | 3.71E-01 | 5.11E-02 | 1.09E-05 | 4.38E-02 | 6.79E-02 |
| S_precentral-sup-part | -4.14E-08 | 9.94E-01 | -1.04E-06 | 7.92E-01 | -4.93E-03 | 1.27E-01 | -4.96E-03 | 1.30E-01 | 2.77E-06 | 5.11E-02 | 1.37E-06 | 5.75E-01 | 2.42E-02 | 4.73E-02 | 3.57E-02 | 6.04E-03 |
| S_suborbital | 6.60E-06 | 1.94E-02 | -2.10E-06 | 5.25E-01 | 8.73E-03 | 3.79E-02 | -4.00E-03 | 4.44E-01 | -2.97E-06 | 5.11E-02 | 1.72E-06 | 3.39E-01 | 1.81E-01 | 5.03E-01 | -1.68E-01 | 5.59E-01 |
| S_subparietal | -4.23E-06 | 1.41E-02 | -6.11E-07 | 7.04E-01 | -6.60E-03 | 1.22E-02 | -8.42E-04 | 6.74E-01 | 2.22E-06 | 1.43E-02 | 2.25E-07 | 7.45E-01 | 7.03E-02 | 2.31E-04 | 1.80E-01 | 1.05E-01 |
| S_temporal_inf | 1.82E-06 | 7.58E-01 | 8.50E-06 | 3.33E-02 | -5.60E-04 | 9.19E-01 | 9.42E-03 | 1.66E-01 | -2.80E-07 | 9.37E-01 | -4.32E-06 | 5.28E-02 | -8.53E-03 | 3.14E-01 | 3.78E-02 | 5.92E-01 |
| S_temporal_sup | -1.37E-06 | 4.17E-01 | 8.28E-06 | 6.02E-10 | -3.17E-03 | 1.39E-01 | 9.84E-03 | 2.03E-07 | 1.00E-06 | 1.87E-01 | -3.33E-06 | 1.07E-05 | 4.98E-02 | 1.64E-03 | 5.94E-02 | 5.04E-02 |
| S_temporal_transverse | 2.28E-06 | 3.49E-01 | 4.66E-06 | 7.18E-02 | 3.25E-03 | 4.03E-01 | 5.86E-03 | 1.66E-01 | -3.26E-07 | 8.92E-01 | -4.47E-07 | 7.45E-01 | -1.76E-01 | 4.26E-01 | -3.33E-02 | 1.14E-02 |

**Summary of with lateralization in diffusion measures and cortical thickness for younger and older adults.**
Displayed is the effect of *SIDE* (positive values indicating a lower left-hemispheric value) with according adjusted P-value (false-discovery-rate).
AD: axial diffusivity, RD: radial diffusivity, FA: fractional anisotropy, CT: cortical thickness.
